# Longitudinal prevalence of neurogenic orthostatic hypotension in the idiopathic Parkinson Progression Marker Initiative (PPMI) cohort

**DOI:** 10.1101/2023.10.16.23297107

**Authors:** Paul Beach, J. Lucas McKay

## Abstract

**Background:** Reported orthostatic hypotension (OH) prevalence in Parkinson disease (PD) varies widely, with few studies evaluating specifically neurogenic-OH (nOH). The ratio of orthostatic heart rate (HR) to systolic blood pressure (SBP) change (Δ) is a valid screening method to stratify nOH/non-nOH but had minimal epidemiologic application.

**Objective:** To estimate the prevalence of nOH and non-nOH in the PPMI using the ΔHR/ΔSBP ratio and examine associations between nOH and various motor and non-motor measures.

**Methods:** Longitudinal orthostatic vitals and motor and non-motor measures were extracted (baseline-month 48). Patients were consensus criteria classified as OH+/-, with ΔHR/ΔSBP sub-classification to nOH (ΔHR/ΔSBP<0.5) or non-nOH (ratio≥0.5). Prevalence was determined across visits. Independent linear mixed models tested associations between nOH/non-nOH and clinical variables.

**Results:** Of N=907 PD with baseline orthostatic vitals, 3.9% and 1.8% exhibited nOH and non-nOH, respectively. Prevalence of nOH/non-nOH increased yearly (P=0.012, chi-square), though with modest magnitude (baseline: 5.6% [95% CI: 4.3-7.3%]; month 48: 8.6% [6.4-11.5%]). nOH patients were older than PD with no OH and nOH was associated with greater impairment of motor and independent functioning than non-nOH/OH-groups. Cognitive function and typical OH symptoms were worse in PD+OH, generally.

**Conclusions:** nOH prevalence was greater than non-nOH in the PPMI early PD cohort, with modest prevalence increase over time. Our findings are consistent with prior studies of larges cohorts that evaluated nOH, specifically. Early PD with nOH were likelier to be older and suffer from greater motor and functional impairment, but OH presence was generally associated with more cognitive impairment.

## Introduction

While orthostatic hypotension (OH) is a common non-motor phenomenon in Parkinson disease (PD) clarity about its prevalence, particularly in early stages, is lacking. Two meta-analyses of mostly cross-sectional studies suggest a pooled estimated prevalence of ∼30%, though the range was ∼10-65%, and additionally provide evidence that OH presence in PD varies as a function of disease duration, age, and dopaminergic dosing^1,2^. Recent prospective studies suggest a prevalence of ∼14-20% in early-stage or de novo PD, with a yearly increase in prevalence thereafter by ∼2%^3,4^. There is evidence of OH in a significant number of individuals at high risk of PD, including isolated REM sleep behavior disorder and pure autonomic failure^5,6^. The variability of prior findings and limited prospective studies of large cohorts indicate a need to enhance understanding of OH longitudinal prevalence particularly in early PD.

Determining the most likely etiology of OH is essential toward prognostication and treatment. The general criteria for OH are a sustained fall in systolic blood pressure (SBP) of ≥20mmHg or ≥10mmHg in diastolic blood pressure (DBP) within 3 minutes of upright posture (head-up tilt or active standing); a more stringent definition of SBP fall ≥30 mmHg or DBP fall ≥15 mmHg may be considered with presence of concurrent supine hypertension (supine blood pressure persistently ≥140/90 mmHg)^7,8^. However, with few exceptions^3,9–11^ most studies thus far examining OH prevalence in PD overlook consideration of whether OH is most likely neurogenic (nOH) or secondary to numerous potential non-neurogenic etiologies (non-nOH; e.g., volume depletion, medication side effects, cardiac pathology). The primary mechanism underlying nOH in PD involves post-ganglionic sympathetic denervation leading to inadequate peripheral vasoconstriction during upright posture^12^. Impaired blood pressure response to a Valsalva maneuver or a blunted supine-to-upright plasma norepinephrine level are two standard methods of differentiating nOH from non-nOH in autonomic testing^13^. However, formal autonomic testing is limited in availability, Valsalva is often confounded by suboptimal expiratory effort or various contraindications to performing the maneuver, and orthostatic catecholamines can be difficult to obtain.

Fortunately, recent work has supported utility of a simple bedside calculation to help differentiate nOH from non-nOH. Specifically, a ratio of orthostatic heart rate change (ΔHR of standing minus supine – beats per minute/bpm) to SBP change (ΔSBP, standing minus supine) less than 0.5 is able to differentiate nOH from non-nOH during tilt testing or active standing with high sensitivity and specificity in populations of patients with OH secondary to synucleinopathy^14,15^. While this calculation is not a definitive means of deducing neurogenicity of OH^16,17^, this ratio provides a clinically practical means of screening for presence of nOH particularly in PD, where baroreflex sympathoneural failure and cardiovagal failure commonly co-occur. Yet the ΔHR/ΔSBP ratio has rarely been utilized in any multi-site, longitudinal analyses examining nOH prevalence in PD.

We thus undertook such an analysis using open-access data from the Parkinson’s Progression Marker Initiative (PPMI), a prospective, multi-site, longitudinal biosample library^18^. Our primary goal was first to determine in this database the overall prevalence of nOH, compared to non-nOH and no OH, and how prevalence changes over time. OH presence in PD is associated with many co-morbidities, including fall risk/injury, faster decline in motor and cognitive function, depression, greater healthcare utilization, reduced quality of life, and a greater mortality risk^19–25^. Multiple studies have utilized the PPMI to examine associations of OH symptoms with these varied co-morbidities, though without confirming concurrent presence of OH^26–32^. This approach is confounded by the large percentage of OH patients who have atypical symptoms or are asymptomatic, as well as the non-specificity of typical OH symptoms assessed in questionnaires^33,34^. In a secondary analysis, we thus also examined the relationship between OH presence, stratified by etiology (nOH or non-nOH), with motor and non-motor measures, assessments of patient daily function, and various autonomic symptoms.

## Methods

### Prevalence of neurogenic and non-neurogenic OH in PPMI data

Patients were initially classified as OH(+/-) at each study observation based on standard criteria: either a drop in systolic blood pressure from supine to standing of 20 mmHg or more, or a drop in diastolic blood pressure from supine to standing 10 mmHg or more. Because we found that a number of patients (≈4.5%) with abnormal diastolic indicators had normal systolic indicators using this formula – and because our orthostatic ratio was based on systolic blood pressure only, we based OH(+/-) classification on systolic criteria only. For the primary analysis we applied discriminative criteria based on the presence of supine hypertension. Patients without supine hypertension were classified as OH (+) based on a drop in systolic blood pressure from supine to standing 20 mmHg or more^7^. Patients with supine hypertension (SBP ≥ 140 mmHg) were classified as OH (+) based on a drop in systolic blood pressure from supine to standing 30 mmHg or more^8^.

Observations were therefore classified OH(+/-) as follows:

Vital Signs at Visit →
if supine SBP < 140 mmHg :

if ΔSBP < 20 mmHg : OH (+)
if ΔSBP ≥ 20 mmHg : OH (-)
if supine SBP ≥ 140 mmHg :

if ΔSBP < 30 mmHg : OH (+)
if ΔSBP ≥ 30 mmHg : OH (+)
Patients classified as OH (+) were further classified as nOH (+)/ non-nOH (+) as follows:

OH (+) →

if ΔHR/ΔSBP < 0.5 : nOH (+)
if ΔHR/ΔSBP ≥ 0.5 : non-nOH (+)

### Data Sources

Data used in the preparation of this article were obtained [on March 6, 2023] from the Parkinson’s Progression Markers Initiative (PPMI) database (www.ppmi-info.org/access-data-specimens/download-data), RRID:SCR_006431. For up-to-date information on the study, visit www.ppmi-info.org.

We used all available data of patients from the “Parkinson’s disease” consensus cohort for whom vital sign measurements were available. We considered data from study visits at Screening, Baseline, and at planned visits at Months 12, 24, 36, and 48. Beginning with N=4061 initial cases, exclusions were as follows: 1) N=34 cases were excluded based on incomplete vital sign data; 2) N=9 cases were excluded based on suspected erroneous measurements of heart rate decelerations from supine to standing of 25 bpm or more. After exclusions, 4018 cases were available for analysis.

All cases were coded as OH(-), nOH(+) or non-nOH(+) based on criteria described above. We noted that among some cases initially coded as non-nOH(+) (N=3), heart rate decelerations, rather than accelerations, were observed between supine and standing. This would suggest that an element of autonomic failure related to baroreflex cardiovagal dysfunction. Based on this, these cases were reclassified as nOH(+).

### Clinical Variables

Several motor and non-motor instruments recorded during PPMI visits were extracted for evaluation of their association with nOH and non-nOH relative to those without OH. Motor function was examined via Part III of the Movement Disorder Society-Unified Parkinson Disease Rating Scale (MDS-UPDRS-III; higher scores = greater motor dysfunction)^35^. Cognitive function testing involved the Montreal Cognitive Assessment (MoCA; higher scores = better cognitive function)^36^. Depression symptoms were assessed using the Geriatric Depression Scale (GDS; higher score = greater depression symptom severity)^37^. The Schwab and England Activities of Daily Living Scale was extracted to evaluate reported ratings of functional independence level (higher = more independence). Last, general autonomic symptoms in the cohort were evaluated with the Scales for Outcomes in Parkinson Disease – Autonomic (SCOPA-AUT; higher scores = more symptomatic)^38^ instrument, with the cardiac domain analyzed independently as well.

Because medication state was not reported for >60% of MDS-UPDRS-III scores, the highest score reported at each visit was used. Disease duration was calculated as the year difference between the enrollment date and either the symptom onset date or the diagnosis date, whichever was earlier. In cases of missing data, covariates were imputed via last observation carry forward if needed. In the case of instruments that were not used at Baseline or Screening (e.g., MoCA), the first available value was used for summarization in demographics tables.

### Statistical analyses

Point estimates and 95% confidence intervals for the prevalence of nOH(+)/non-nOH(+)/OH(-) across visits were calculated using Wilson score formulae^39^. A chi-squared test was used to evaluate the statistical significance of overall changes in crude prevalence across time. Linear mixed models (lmerTest::lmer) were used to identify differences in clinical and demographic covariates between patients with nOH(+)/non-nOH(+)/OH(-) after controlling for fixed effects of age and sex and random effects for patients. As age and sex were used as covariates in other models, differences in patient age across groups and frequency of female sex across groups were assessed with a linear mixed model a random effect for patient and with a logistic regression model, respectively. Missing covariate data were imputed via last observation carry forward prior to entry in linear mixed models. Statistical tests were performed in R version 4.2.1 and evaluated at alpha=0.05.

### Additional analyses

To evaluate sensitivity to the specific criteria used for OH in the primary analyses, we iterated analyses of prevalence estimates using the less stringent OH criterion of SBP fall of 20 mmHg or more (see Supplemental Information).

## Results

### Clinical and demographic variables

Clinical and demographic variables are summarized and stratified by nOH(+)/non-nOH(+)/OH(-) at the first available study visit in Table 1. For items that were not taken at baseline or screening, the earliest values are presented. Supplementary Table S1 shows these variables without the more conservative definition of OH with presence of concurrent supine hypertension.

**Table 1.** Demographic and clinical features of the study sample at the first available visit.

|  | nOH(+) | non-nOH(+) | OH(-) |
| --- | --- | --- | --- |
| N \| % | 35 \| 3.8% | 16 \| 1.7% | 873 \| 94.5% |
| Age, y |  |  |  |
| Mean (SD) | 68 (9) | 61 (13) | 62 (10) |
| Range | 39 – 82 | 29 – 73 | 30 – 85 |
| Sex (N, %) |  |  |  |
| Female | 13 (37%) | 6 (37%) | 347 (40%) |
| Male | 22 (63%) | 10 (63%) | 526 (60%) |
| MDS-UPDRS-III |  |  |  |
| Mean (SD) | 26.2 (12.3) | 23.5 (11.3) | 21.8 (9.9)<sup>a</sup> |
| Range | 5 – 55 | 5 – 39 | 4 – 71 |
| Duration, y |  |  |  |
| Mean (SD) | 2.9 (2.8) | 4.6 (5.4) | 2.9 (2.8) |
| Range | 0.3 – 13.7 | 0.4 – 20.1 | 0.2 – 22.9 |
| Supine Hypertension |  |  |  |
| Absent | 19 (54%) | 14 (88%) | 623 (71%) |
| Present | 16 (46%) | 2 (12%) | 250 (29%) |
| Supine Systolic BP, mmHg |  |  |  |
| Mean (SD) | 139.5 (19.8) | 130.6 (11.8) | 130.2 (17.3) |
| Range | 110 – 212 | 110 – 159 | 82 – 200 |
| Supine Diastolic BP, mmHg |  |  |  |
| Mean (SD) | 79.5 (11.8) | 79.0 (7.6) | 77.5 (10.4) |
| Range | 54 – 109 | 61 – 94 | 35 – 121 |
| Supine Heart Rate, bpm |  |  |  |
| Mean (SD) | 69.0 (11.3) | 66.0 (11.0) | 67.7 (10.8) |
| Range | 45 – 89 | 50 – 84 | 37 – 103 |
| Delta SBP |  |  |  |
| Mean (SD) | -33.6 (9.7) | -26.4 (8.1) | -2.1 (10.2) |
| Range | -72 – -20 | -49 – -20 | -29 – 51 |
| Delta HR |  |  |  |
| Mean (SD) | 6.6 (4.9) | 18.3 (5.6) | 7.1 (7.1) |
| Range | -3 – 17 | 11 – 29 | -18 – 31 |
| Delta HR / Delta SBP |  |  |  |
| Mean (SD) | 0.21 (0.13) | 0.71 (0.21) | 1.76 (2.80)<sup>b</sup> |
| Range | 0.00 – 0.46 | 0.50 – 1.15 | 0.00 – 23.00 |

Abbreviations: MDS-UPDRS-III, Unified Parkinson's Disease Rating Scale Part III, Movement Disorders Society Revision; SBP, Systolic Blood Pressure; DBP, Diastolic Blood Pressure; HR, Heart Rate. <sup>a</sup>N=862. <sup>b</sup>N=865; this ratio was 0/0 for N=8 cases.

### OH prevalence over time

The prevalence of all-cause OH among the study sample was not constant across visits (P=0.012, chi-square), but the numerical changes in prevalence were small, from 5.6% [95% CI: 4.3-7.3%] at baseline visit to 8.6% [6.4-11.5%] at month 48. No significant prevalence changes year-over-year were identified in post-hoc chi-squared tests. Among OH(+) patients, nOH was 3.4 times more prevalent than non-nOH on average. Prevalence of nOH(+)/non-nOH(+)/OH(-)stratified by study visit is presented in Table 2. Prevalence data whereby less stringent criteria for defining OH were applied (i.e., not accounting for presence of supine hypertension) is in Supplementary Table S2. There was an average increase of 2.7% across all visits for patients meeting less stringent nOH criteria and only an average increase of 0.8% for patients meeting non-nOH criteria.

**Table 2.**
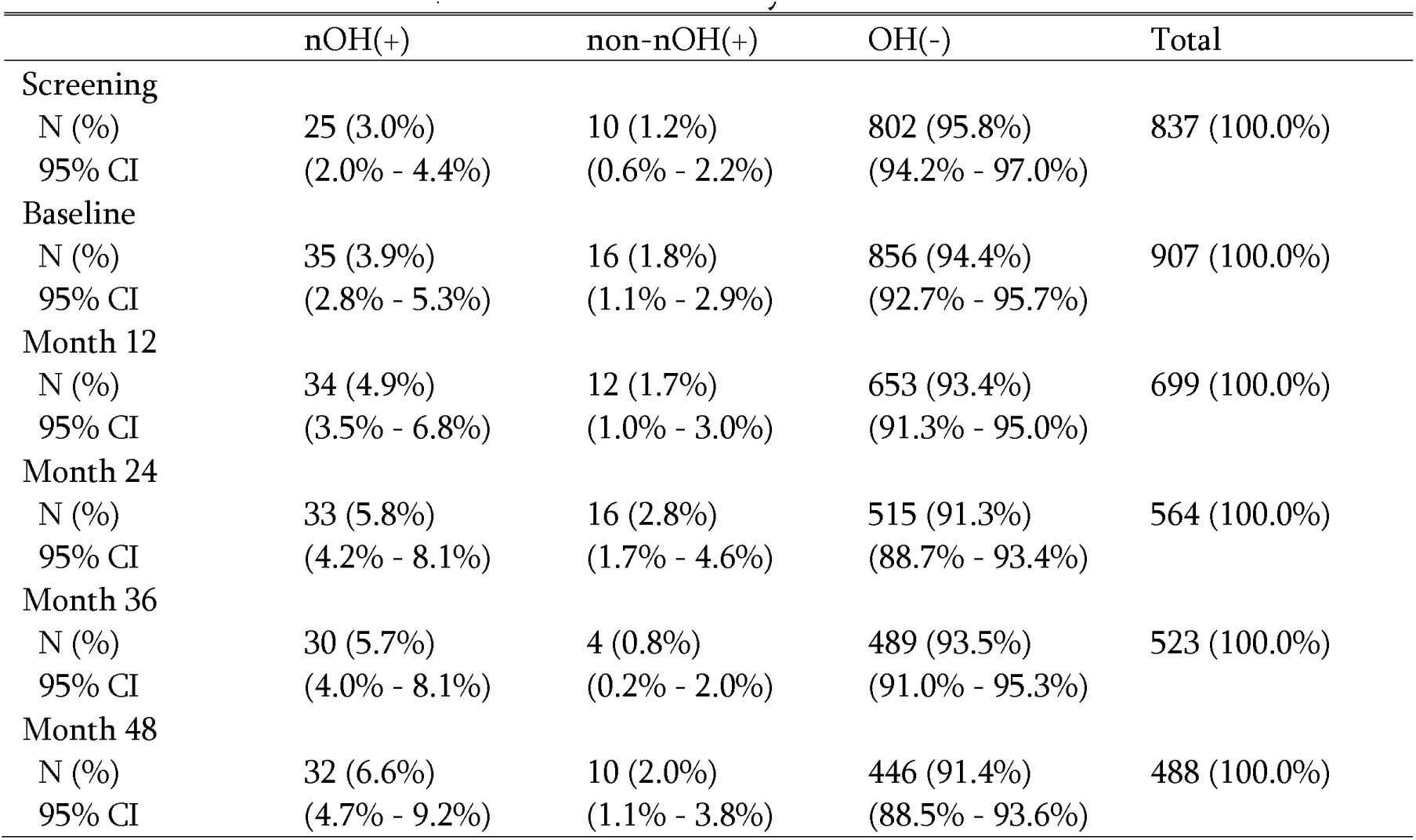
Prevalence of nOH/non-nOH across study visits.

### Clinical variables associated with nOH/non-nOH

While controlling for effects of age and gender, linear mixed models identified: significantly increased age (5.3 year difference in average age) in nOH(+) compared to OH(-); significantly increased MDS-UPDRS-III score (5.1 points) in nOH(+) compared to OH(-); significant decreases in MoCA score in nOH(+) and non-nOH(+) compared to OH(-) of 1.3 points and 0.8 points, respectively; significant decreases in Schwab and England score in nOH(+) compared to non-nOH(+) and OH(-) of 3.7 points and 4.0 points, respectively; and significant increases in SCOPA-AUT cardiac subscore in nOH(+) and non-nOH(+) compared to OH(-) of 0.6 and 0.3 points, respectively. There were no differences amongst groups for GDS scores or overall SCOPA-AUT scores. No differences in gender were observed across groups. These data are summarized in Table 3.

**Table 3.**
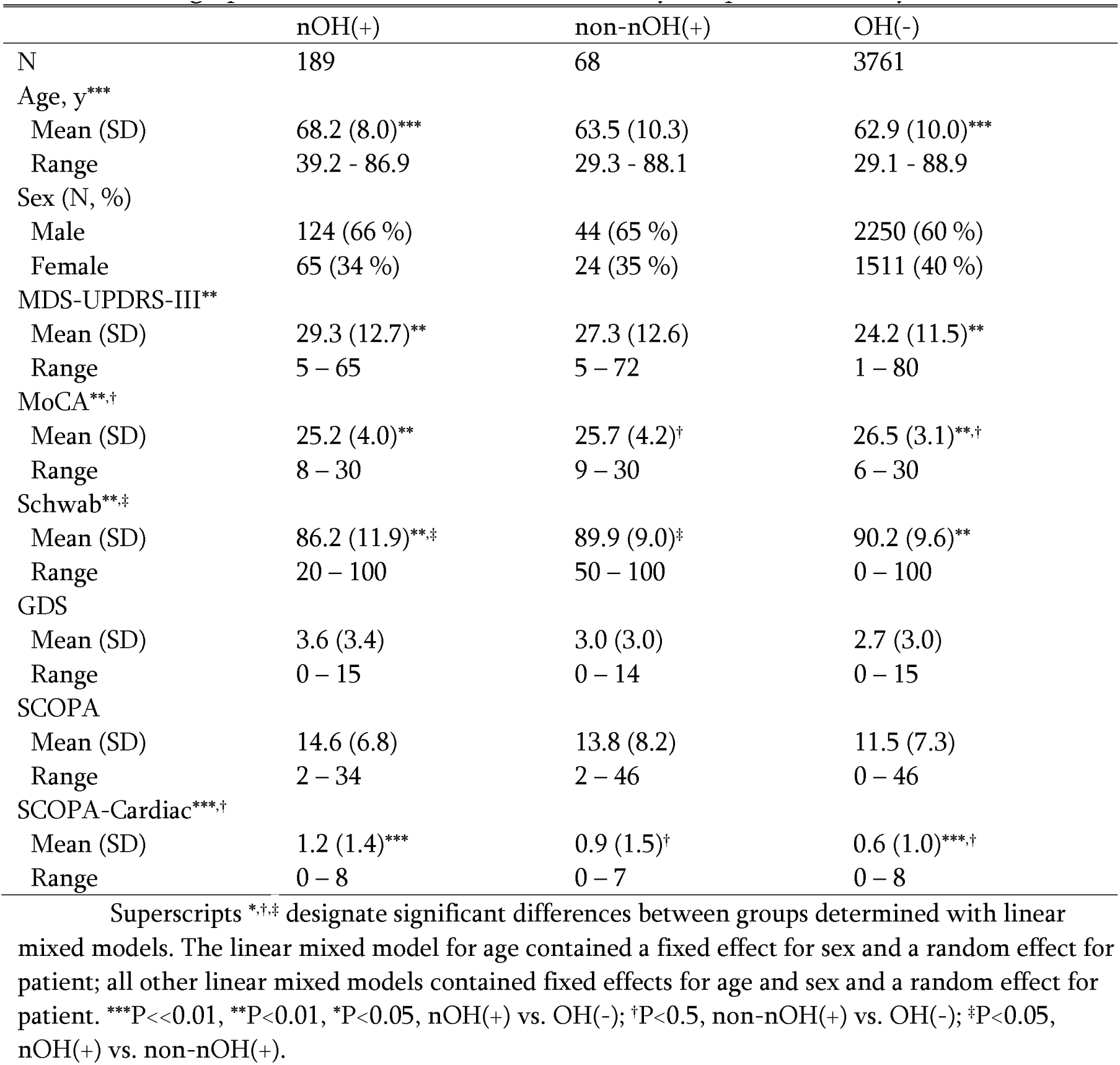
Demographic and clinical features of the study sample across study visits.

## Discussion

The prevalence of nOH in early PD, relative to non-neurogenic forms of OH (non-nOH), changes in prevalence over the course of early PD, and associations of nOH with various motor, non-motor, and independent function assessments have been minimally investigated. We applied the ratio of ΔHR/ΔSBP to orthostatic vitals measured in PD patients participating in the PPMI to discriminate between nOH/non-nOH and estimate their respective longitudinal prevalence. A more conservative criterion for determination of OH was used in the primary analysis in the setting of supine hypertension. We subsequently examined the relationship between nOH presence and symptoms of OH as well as motor and non-motor measures and activities of daily living, relative to patients without OH and those with non-nOH. We found that: first, nOH in the PPMI cohort is far less prevalent than reported in most prior studies examining all-cause OH in PD over a wide range of disease durations, but is more common than non-nOH; second, there was an overall small increase in nOH prevalence over the available 5-year time period. We additionally found certain demographic, motor, and non-motor measures differed between those with nOH and without OH, with few significant differences therein between nOH from non-nOH.

At the baseline visit, prevalence of nOH in the PPMI database was 3.9%, with an increase to 6.6% by month 48. All-cause OH (i.e., nOH + non-nOH) was only slightly higher at baseline, at 5.7%, increasing to 8.8% at the end of the surveyed period. A less stringent analysis (see Supplementary materials), whereby OH designation did not take supine hypertension into account, added an average of less than 3% to our prevalence estimates of nOH and had virtually no impact on all-cause OH estimates. Our findings contrast with most prior studies examining general (all-cause) OH occurrence in PD, which have suggested a much higher prevalence. For example, a pooled estimated prevalence was near 30% in two meta-analyses^1,2^. However, the variability of prevalence amongst individual studies included in these analyses was wide, ranging from as little as ∼4%^9^ to as high as ∼60%^3,40^. This variability is additionally reflected in the few studies that have examined longitudinal prevalence, with yearly increases ranging from ∼2%^3,9^ to ∼15%^41^. These inconsistencies are, in part, likely a reflection of commonly low, but highly varied sample sizes^1,2^. Indeed, one recent study that utilized PPMI data, by Chen et al^4^., found a 13% prevalence of OH at the baseline visit, but had 408 fewer patients included at the time of that study compared to our baseline count. Prior studies have additionally differed in timing of BP measurement after standing, number of BP measures, OH definition including accounting (or not) for supine hypertension, and discrimination (or not) of OH etiology. An additional possible reason for reduced prevalence here, in comparison to other studies, are the criteria for PPMI recruitment, which in part mandates exclusion of individuals of a disease duration greater than two years. Additionally, a diagnosis of PD with early autonomic failure may be confused with parkinsonian-variant multiple system atrophy^42^. These latter two factors may have prevented PD patients with early OH from PPMI participation. Nevertheless, our findings correspond very well with one of only three other studies that specifically investigated prevalence of nOH in PD. Baschieri and colleagues^9^, as part of a prospective study of 105 well-characterized, early and mostly de novo PD patients utilized formal autonomic testing, including head-up tilt, and accounting for supine hypertension to both diagnose and discriminate nOH vs non-nOH. They found nOH prevalence in their sample to be 3.8% at baseline, increasing to 7.6% after 16 months. Two other studies have utilized the ΔHR/ΔSBP ratio to classify nOH. Yoo et al.^10^ recently reported that, amongst 267 de novo PD patients, 17.2% had nOH (based on the ΔHR/ΔSBP ratio) during head-up tilt at a baseline visit, increasing to 31.2% on follow up (∼3 years). While the authors corrected for supine hypertension, there are inconsistencies regarding OH prevalence in the sample making it unclear whether the latter study utilized a definition of sustained blood pressure fall throughout tilt testing. Dommershuijsen and colleagues^11^ applied the nOH ratio to orthostatic vitals taken as part of a population-based study (>6,000 individuals) that included 62 individuals with PD, finding a baseline nOH prevalence of 21% the PD group, which was also older and of a longer disease duration than those in the PPMI and Baschieri sample. Indeed, multiple prior investigations of OH/nOH prevalence, including our own findings, suggest particularly age and probably disease duration play a role in the occurrence of OH in PD^2–4,10,40^. Thus, alongside variations in sample sizes, the range of ages and disease durations included in prior studies are also likely significant factors in explaining differences between our baseline and longitudinal results and those of prior studies.

Aside from informing treatment on an individual level, discriminating between nOH vs non-nOH, is informative regarding prognosis. OH in PD has been found associated with many co-morbidities including faster motor progression and risk of falls, greater depression and cognitive decline, increased mortality, increased financial burden, reduced functional independence, and reduced quality of life^19–22,24,25,41^. We replicated some of these findings in the current study, particularly with respect to individuals meeting nOH criteria. Regarding motor dysfunction, our nOH sample had higher MDS-UPDRS III scores than those without OH but did not differ from those with non-nOH. This general finding has been reported many times, but it remains unclear whether this association reflects a more aggressive PD phenotype or a shared tendency toward inactivity/deconditioning, which can worsen OH and motor impairment^43–48^. Both nOH and non-nOH performed worse on cognitive testing than those without OH, though the average magnitude of score differences was not large. This finding is consistent with the presence of OH generally conferring a greater risk of cognitive decline not just in PD^20,49–51^, but even in population samples of individuals at mid-life^52^, perhaps through chronic non-specific hypoxia induced neurodegeneration^51^. One recent study did find presence of cognitive decline specifically amongst nOH, compared to non-nOH^53^. However, these authors used formal neuropsychological testing, compared to use of a cognitive screening (MoCA) in the PPMI. We additionally found that nOH specifically was associated with reduced functional independence, compared to individuals with non-nOH and those without OH. This is consistent with prior studies of self or caregiver rated functional independence levels in patients nOH^54^. In contrast to prior work, we found no statistically-significant associations between nOH/non-nOH and symptoms of depression. This may relate to many prior studies finding this association with respect to autonomic symptom report, rather than actual nOH/non-nOH diagnosis, or use of alternative scales to the GDS^55^, which is utilized in the PPMI^24,26,32,56^.

Several reports now indicate a lack of reliability for symptom report to reflect both OH occurrence and severity, seriously calling into question the validity of relying on symptom report as an adequate screen for OH^9,25,33,34,49^. We found no general relationship between number of reported autonomic symptoms (total SCOPA-AUT scores) and OH presence. However, typical OH symptoms, as surveyed by the cardiac domain of the SCOPA-AUT, were more common in those with nOH and non-nOH, compared to those without OH. This finding would, in part, support face validity of the cardiac domain of the SCOPA-AUT, at least from the standpoint of patient recall of past symptoms.

The orthostatic ΔHR/ΔSBP ratio is a practical clinical tool to screen for nOH given the common difficulty of obtaining formal autonomic function testing. While shown to be highly sensitive to nOH presence in patients with synucleinopathies during head-up tilt^14,15^, the ratio as a screening measure can be susceptible to reduced sensitivity during active standing due to inherent differences between these orthostatic challenges^15^. There have also been reports of false negative findings in validation studies using head-up tilt, with speculation that the ratio is best utilized in patients with predominantly peripheral forms of autonomic degeneration, such as PD, dementia with Lewy bodies, or pure autonomic failure^17^; its use in the PPMI thus seems especially valid. Potential limitations of the ratio aside, a significant strength of this study is the use of this novel screening tool in a large, well-characterized longitudinal PD cohort. We have additionally replicated similar prevalence numbers to those of Baschieri and colleagues, who used gold-standard autonomic measures to diagnose OH and discriminate between nOH/non-nOH in a, comparatively speaking, moderately sized early PD cohort. A limitation to our approach of using PPMI data involves the nature of the PPMI cohort, which currently skews toward a low disease duration and younger in age. Re-evaluation of nOH/non-nOH prevalence over more time in this cohort will help address this limitation. Another limitation relates to the orthostatic challenge utilized in the PPMI, which involves a single standing measure. Multiple measures (e.g., at 1 and 3 minutes), or for a longer period, would ensure capture of a sustained fall in BP as per OH criteria. Longer measures would also help delineate the prevalence of delayed OH in PD, which occurs in earlier stages of autonomic failure, yet still confers a high long-term risks and does tend to progress in severity over time^57,58^. Also, our analysis did not control for use of anti-hypertensive or anti-OH medication, which may have inflated prevalence numbers somewhat. Though, of note, the PPMI does not indicate whether these medications were taken around the time of vitals testing; we additionally found many motor evaluations did not include indication of OFF or ON state of anti-parkinsonian medication. Last, some cases were excluded from nOH designation due to meeting OH criteria through diastolic BP fall only, or due to presence of supine hypertension. However, based on case counts and reviewing prevalence data without correcting for supine hypertension, we see that this minimally impacted our overall prevalence numbers (see supplementary data).

In conclusion, this study utilized an easily applicable screening tool, the ΔHR/ΔSBP ratio, to estimate the longitudinal prevalence of nOH and non-nOH in a large, well-characterized population of early PD patients, the PPMI cohort. We found that nOH was more common than non-nOH in the PPMI sample. On the one hand, our pooled prevalence estimates of nOH and non-nOH, as well as longitudinal changes therein, is much lower than findings reported in many prior studies evaluating all-cause OH in much smaller samples of patients of varying disease durations and ages. However, our results are very similar to recent work evaluating nOH prevalence using gold standard autonomic testing methods in early PD^9^. In addition, we replicated prior findings associating all-cause OH with increased age, greater motor and cognitive impairment, and reduced functional independence. Lastly, we found OH symptom report increased in nOH and non-nOH compared to no OH. These findings support the use of the ΔHR/ΔSBP ratio as a means of screening for nOH in PD, as well as the need to diligently screen for OH generally in PD given co-morbid risks. Our findings additionally suggest caution in attempting to estimate OH prevalence with small sample sizes.

## Supporting information

PPMI_OH_supplementary-tables

## Data Availability

All data are available from the data source: www.ppmi-info.org.

## Supplemental Information

### Simplified OH criterion

We iterated the main analyses with a simplified criterion for discriminating nOH(+) from non-nOH(+) that was independent of the presence of supine hypertension. Observations were therefore classified OH(+/-) as follows:

Vital Signs at Visit →

if ΔSBP < 20 mmHg : OH (+)
if ΔSBP ≥ 20 mmHg : OH (-)
Patients classified as OH (+) were further classified as nOH (+)/ non-nOH (+) as follows: OH (+) →

if ΔHR/ΔSBP < 0.5 : nOH (+)
if ΔHR/ΔSBP ≥ 0.5 : non-nOH (+)

Qualitatively similar results were obtained using either criterion. Although as expected prevalence estimates were somewhat higher than in the main text, overall OH prevalence estimates never exceeded 31.1% at any given timepoint. Tables S1 and S2 replicate the information presented in Tables 1 and 2 of the main text using this simplified OH criterion.

### Clinical and demographic variables using simplified OH criterion

Clinical and demographic variables are summarized stratified by nOH(+)/non-nOH(+)/OH(-) at the first available study visit are summarized in Table S1. For items that were not taken at baseline or screening, the earliest values are presented.

### Prevalence over time using simplified OH criterion

The prevalence of all-cause OH among the study sample was not constant across visits (P=0.009, chi-square), but the numerical changes in prevalence were small, from 7.0% at screening to 11.4% at month 48. No significant prevalence differences over years were identified in chi-squared tests applied post-hoc on year-to-year changes. OH prevalence stratified by study visit is presented in Table S2.

### Covariate selection

The covariates selected for analysis for association with nOH(+)/non-nOH(+)/OH(-) were Age, Sex, MDS-UPDRS-III, MoCA score, Schwab and England Score, GDS, SCOPA, and SCOPA Cardiac subscale. PD duration was not entered into analyses because of total collinearity with Age.

**Table S1.**
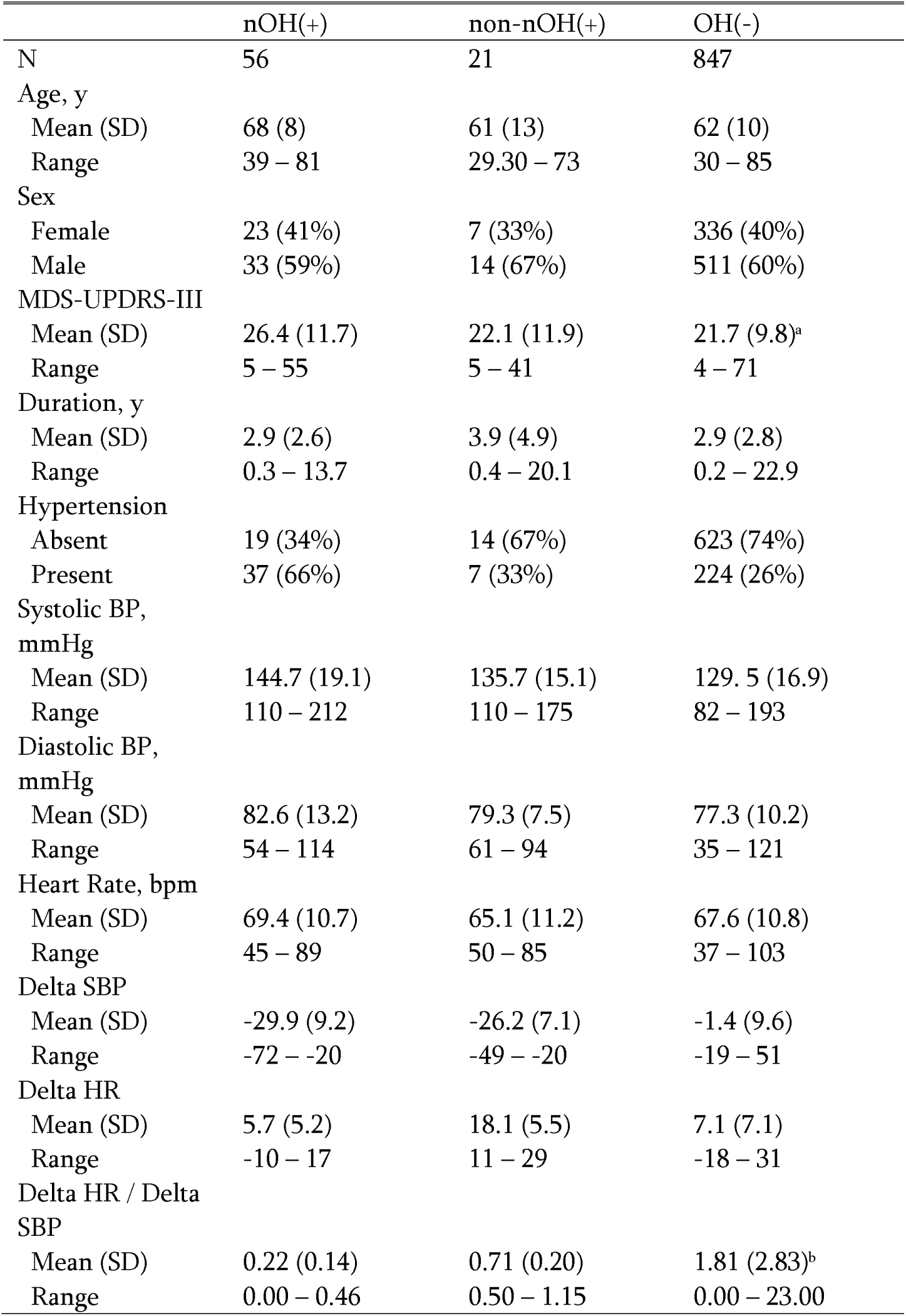
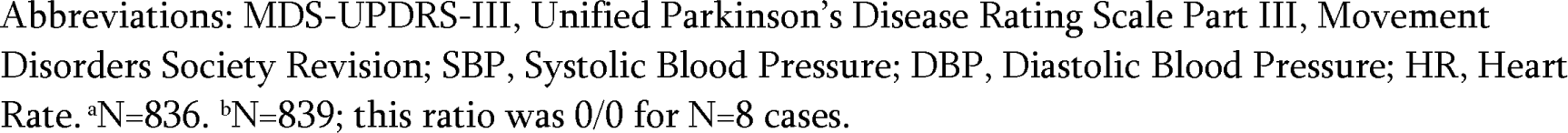
Demographic and clinical features of the study sample at the first available visit, using a simplified OH criterion.

**Table S2.**
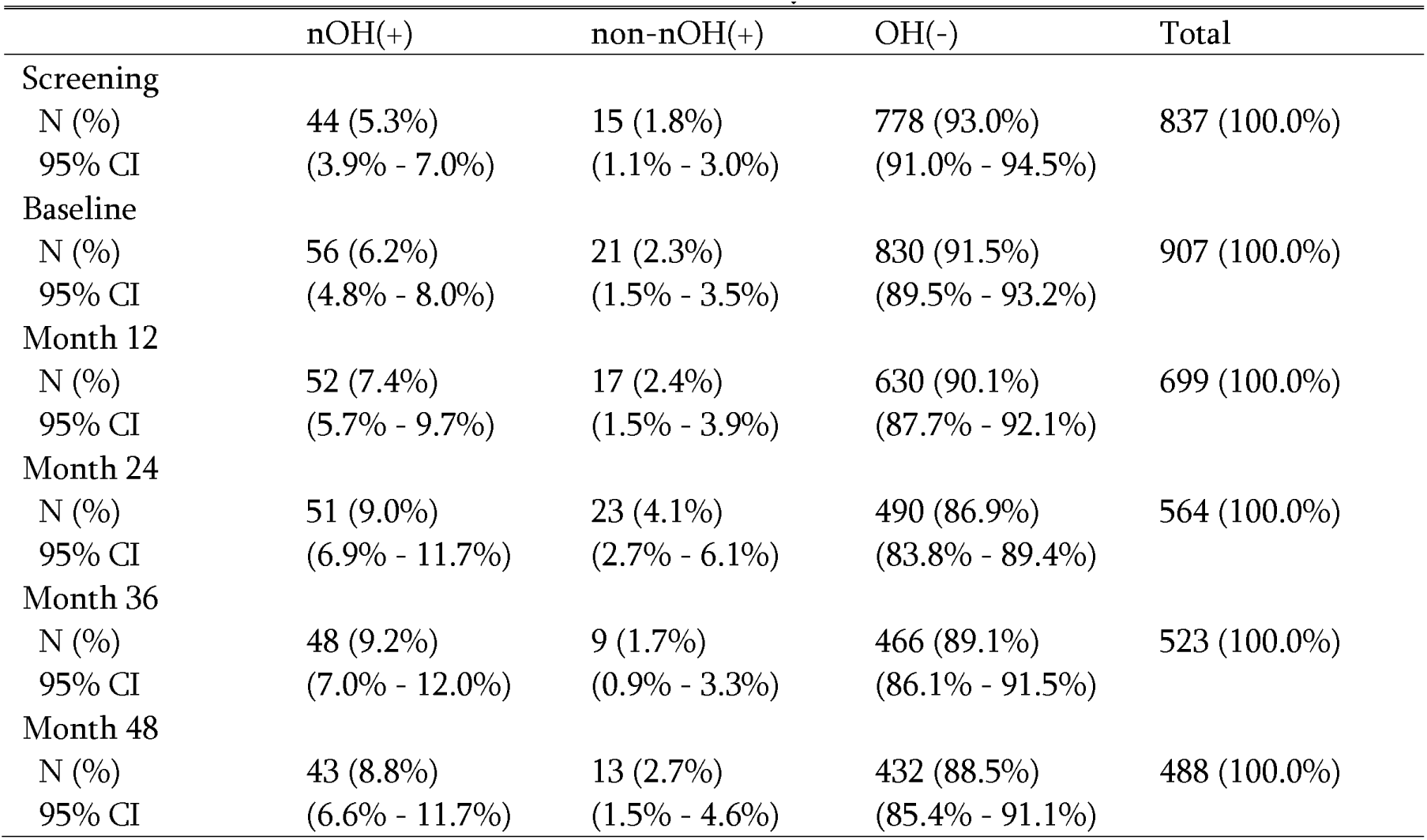
Prevalence of nOH/non-nOH across study visits.

