## Supplementary material for "Longitudinal prevalence of neurogenic orthostatic hypotension in the idiopathic Parkinson Progression Marker Initiative (PPMI) cohort": PPMI_OH_supplementary-tables

| Table S1. Demographic and clinical features of the study sample at the first available visit, using a simplified OH criterion. | | | |
| --- | --- | --- | --- |
|  | nOH(+) | non-nOH(+) | OH(-) |
| N | 56 | 21 | 847 |
| Age, y |  |  |  |
| Mean (SD) | 68 (8) | 61 (13) | 62 (10) |
| Range | 39 – 81 | 29.30 – 73 | 30 – 85 |
| Sex |  |  |  |
| Female | 23 (41%) | 7 (33%) | 336 (40%) |
| Male | 33 (59%) | 14 (67%) | 511 (60%) |
| MDS-UPDRS-III |  |  |  |
| Mean (SD) | 26.4 (11.7) | 22.1 (11.9) | 21.7 (9.8)^a^ |
| Range | 5 – 55 | 5 – 41 | 4 – 71 |
| Duration, y |  |  |  |
| Mean (SD) | 2.9 (2.6) | 3.9 (4.9) | 2.9 (2.8) |
| Range | 0.3 – 13.7 | 0.4 – 20.1 | 0.2 – 22.9 |
| Hypertension |  |  |  |
| Absent | 19 (34%) | 14 (67%) | 623 (74%) |
| Present | 37 (66%) | 7 (33%) | 224 (26%) |
| Systolic BP, mmHg |  |  |  |
| Mean (SD) | 144.7 (19.1) | 135.7 (15.1) | 129. 5 (16.9) |
| Range | 110 – 212 | 110 – 175 | 82 – 193 |
| Diastolic BP, mmHg |  |  |  |
| Mean (SD) | 82.6 (13.2) | 79.3 (7.5) | 77.3 (10.2) |
| Range | 54 – 114 | 61 – 94 | 35 – 121 |
| Heart Rate, bpm |  |  |  |
| Mean (SD) | 69.4 (10.7) | 65.1 (11.2) | 67.6 (10.8) |
| Range | 45 – 89 | 50 – 85 | 37 – 103 |
| Delta SBP |  |  |  |
| Mean (SD) | -29.9 (9.2) | -26.2 (7.1) | -1.4 (9.6) |
| Range | -72 – -20 | -49 – -20 | -19 – 51 |
| Delta HR |  |  |  |
| Mean (SD) | 5.7 (5.2) | 18.1 (5.5) | 7.1 (7.1) |
| Range | -10 – 17 | 11 – 29 | -18 – 31 |
| Delta HR / Delta SBP |  |  |  |
| Mean (SD) | 0.22 (0.14) | 0.71 (0.20) | 1.81 (2.83)^b^ |
| Range | 0.00 – 0.46 | 0.50 – 1.15 | 0.00 – 23.00 |
| Abbreviations: MDS-UPDRS-III, Unified Parkinson’s Disease Rating Scale Part III, Movement Disorders Society Revision; SBP, Systolic Blood Pressure; DBP, Diastolic Blood Pressure; HR, Heart Rate. ^a^N=836. ^b^N=839; this ratio was 0/0 for N=8 cases. | | | |

| Table S2. Prevalence of nOH/non-nOH across study visits. | | | | |
| --- | --- | --- | --- | --- |
|  | nOH(+) | non-nOH(+) | OH(-) | Total |
| Screening |  |  |  |  |
| N (%) | 44 (5.3%) | 15 (1.8%) | 778 (93.0%) | 837 (100.0%) |
| 95% CI | (3.9% - 7.0%) | (1.1% - 3.0%) | (91.0% - 94.5%) |  |
| Baseline |  |  |  |  |
| N (%) | 56 (6.2%) | 21 (2.3%) | 830 (91.5%) | 907 (100.0%) |
| 95% CI | (4.8% - 8.0%) | (1.5% - 3.5%) | (89.5% - 93.2%) |  |
| Month 12 |  |  |  |  |
| N (%) | 52 (7.4%) | 17 (2.4%) | 630 (90.1%) | 699 (100.0%) |
| 95% CI | (5.7% - 9.7%) | (1.5% - 3.9%) | (87.7% - 92.1%) |  |
| Month 24 |  |  |  |  |
| N (%) | 51 (9.0%) | 23 (4.1%) | 490 (86.9%) | 564 (100.0%) |
| 95% CI | (6.9% - 11.7%) | (2.7% - 6.1%) | (83.8% - 89.4%) |  |
| Month 36 |  |  |  |  |
| N (%) | 48 (9.2%) | 9 (1.7%) | 466 (89.1%) | 523 (100.0%) |
| 95% CI | (7.0% - 12.0%) | (0.9% - 3.3%) | (86.1% - 91.5%) |  |
| Month 48 |  |  |  |  |
| N (%) | 43 (8.8%) | 13 (2.7%) | 432 (88.5%) | 488 (100.0%) |
| 95% CI | (6.6% - 11.7%) | (1.5% - 4.6%) | (85.4% - 91.1%) |  |
